# Hospital at home: a systematic review of how medication management is conceptualised, described and implemented in practice – a study protocol

**DOI:** 10.1101/2022.05.10.22274917

**Authors:** Sophie McGlen, Daniel Lasserson, Clare Crowley, Zahra AL Qamariat, Rosemary H Lim

## Abstract

**Introduction:** Hospital at Home (H@H) is a method of healthcare delivery, where hospital level interventions are conducted in the patient’s usual place of residence, offering an alternative to hospital admission. This often includes the ability to perform point of care diagnostics and treat conditions using a range of treatments traditionally associated with hospital admission, including intravenous medicines and oxygen. H@H services have been established worldwide but there is a wide variation in definition and delivery models and currently no documented evidence supporting the delivery of medicines and medicines management within the H@H model. Therefore, this study aims to 1) describe how medication management in H@H is conceptulised, 2) describe and identify key components of medication management in H@H and 3) describe and identify variability in the implementation of medication management services within H@H models

**Methods and Analysis:** We will search a range of databases (PubMed, Medline, Embase, CINAHL), publicly accessible documents and expert recommendations. Studies, reports and policy documents published between 1^st^ January 2000 and 31^st^ January 2022 will be included. Two independent reviewers will 1) screen and select studies based on *a priori* inclusion/exclusion, 2) conduct quality assessment using the Mixed Methods Appraisal Tool on included studies and 3) extract data. Inductive thematic analysis (objectives 1 and 2), the SEIPS 2.0 model (objective 2) and the Consolidated Framework for Implementation Research (objective 3) will be used to synthesise data.

**Ethics and dissemination:** This systematic review will use secondary data sources from published documents, and as such research ethical approval was not required. We will disseminate the findings of this study in a peer-reviewed journal and national/international conference(s).

**PROSPERO registration number:** CRD42022300691

**Strengths and limitations of this study:**

- The search will be performed on a comprehensive range of databases and relevant information sources to capture a global perspective relating to Hospital at Home (H@H).
- A wide range of search terms will be used in our search, however due to the changing nature of H@H, some terms may be unintentionally missed.
- Only published literature will be synthesized in this systematic review, and it is recognised that in a rapidly changing environment not all practices will be captured in written form.
- A combination of thematic synthesis, a work systems model (Systems Engineering Initiative for Patient Safety 2.0) and the Consolidated Framework for Implementation Research will be used to provide a system-level analysis of studies.
- The study team consists of four pharmacists from a range of practice and research backgrounds and a H@H consultant, therefore, will benefit from different perspectives on the topic area.

## Introduction

The way in which hospital beds are used is constantly changing. Demand on hospitals in developed countries [1] has been influenced by a growing, ageing population with more chronic health problems and rapid advances in healthcare technology [2,3]. To meet this rising demand within a constrained funding envelope, inpatient healthcare delivery has also evolved. These changes include a move to day-case [4] and ambulatory care pathways (care delivered without a traditional inpatient bedbase) [5], strategies to avoid admission to hospital [6], rapid discharge interventions including to intermediate care facilities (such as respite, reablement or rehabilitation services outside of an acute hospital) [7] and centralisation of services [8]. The number of inpatient beds per population varies greatly globally, even in developed countries [9], driving innovation for alternative healthcare technologies. Despite the introduction of these changes, the hospital system continues to experience staffing shortages, insufficient funding, inadequate space and deteriorating estate, outdated and insufficient provision of IT, falling bed numbers, long waits and waiting lists.

The COVID-19 pandemic has further highlighted and exacerbated the challenges that global healthcare has been facing. Staff were redeployed and procedures cancelled [10], creating a backlog of care and impacting on healthcare professional training [11]. At the same time, COVID-19 highlighted inequities in care provision [12] and the importance of investing in staff welfare [9,13]. As the global health systems face unprecedented pressures, there is recognition that the healthcare system needs to learn, be able to flex and adapt so that it can respond effectively now and in the future. The COVID-19 context has therefore created opportunities to explore and accelerate improved and innovative care models that release inpatient bed capacity in a safe and effective manner [14]. Examples include enlisting the help of the private sector, rapidly evolving digital health technology [15] and providing hospital level care at home [16].

The recent challenges have accelerated the emergence and development of Hospital at Home (H@H) programmes across the globe [12]. H@H is a treatment model that delivers acute healthcare treatment in the patient’s usual place of residence. This model of care has existed in isolated pockets worldwide for several decades, however there is significant variation between how they are organised and delivered. Studies have also demonstrated similar mortality rates between those admitted to hospital and those treated at home [17]. The additional benefits include a reduction in those needing long term residential care following acute illness and reduction in delirium [17]. The key services that unite these differing models is that they provide some element of acute care that under a traditional healthcare model would have been administered as an inpatient in an acute hospital setting.

There is no internationally recognised definition of H@H, however within the UK, the UK Hospital at Home society released the following consensus statement [18]:

> *“Hospital at home provides intensive hospital level care for acute conditions that would normally require an acute hospital bed, in a patient’s home for a short episode through multidisciplinary healthcare teams”*.

Other services from around the world have similar explanations of their services describing a hospital level intervention. In the UK these have often been termed virtual wards, and have received increasing support from the UK government through NHS England to roll out more comprehensive virtual wards delivering H@H interventions [19]. In the USA, an Acute Hospital Care at Home waiver announced during the pandemic [20] has allowed patients to receive at home care and providers gain reimbursement through their insurance programmes in some parts of the country.

Central to the delivery of the hospital level care is the ability to deliver medications within the patient’s home. This is critical in order to act in a timely fashion to the patient’s condition, clinical assessment and point of care diagnostics. This includes medications traditionally restricted to a hospital setting such as intravenous therapies and oxygen. Currently there is no evidence supporting the delivery of medicines and medicines management within the H@H model. With a wide variation in definition and delivery models of H@H services worldwide, describing these will assist with the understanding of services to support the development of future H@H programmes in the current climate.

## Aim and objectives

Therefore, this systematic review aims to answer how medication management within H@H services established worldwide is conceptulised, described and implemented in practice. The objectives of this review are:

- To describe how medication management in H@H is conceptulised
- To describe and identify key components of medication management in H@H
- To describe and identify variability in the implementation of medication management services within H@H models

## Method

### Protocol and registration

This systemic review is registered in the International Prospective Register of Systematic Reviews (PROSPERO), registration number: CRD42022300691. We report this protocol following the Preferred Reporting Items for Systematic Review and Meta-Analysis Protocols (PRISMA-P) statements [21].

### Eligibility criteria

We will include published studies of any study design, reports and policy documents written on and between 1^st^ January 2000 (this was when the concept of H@H was first introduced but the concept has changed over the years) and 31^st^ January 2022 in the English language that meets one or more of the review objectives. Criteria specifically related to PI(E)COS are:

- Participants/population: anyone receiving hospital care at their usual place of residence.
- Intervention(s), exposure(s): medication management in a H@H setting.
- Comparator: where relevant e.g. trial of different medication management approaches
- Outcomes: any
- Study design: any

### Information sources and search strategy

The following international electronic databases will be used: Pubmed, Cochrane, grey literature, Web of Science, Cinahl, Medline, organisation websites/resources such as NHS England and Hospital at Home User Group, references of references and experts’ recommendations. These keywords will be used to search studies: hospital in the home, hospital at home, hospital@home, virtual ward, pharmacist, pharmacy, medication, drug delivery. Searches will use Index Terms unique to each database and a combination of Boolean (AND/OR) keywords, as relevant.

### Study selection

The search results will be collated on a web based systematic review tool (https://www.rayyan.ai/), and duplicates removed. Independent screening of titles and abstracts will be conducted by two researchers applying pre-specified inclusion and exclusion criteria. Where there are disagreements about eligibility of papers, a third reviewer will assess the paper a consensus method used to determine inclusion. Full-text articles of remaining references will then be obtained and screened independently by two researchers using the same inclusion/exclusion criteria and any disagreements will be resolved by discussion to achieve consensus.

### Data extraction

The following information will be extracted: authors, year of publication, country where study was conducted (empirical articles) or researchers were based (non-empirical articles), aim of article, descriptions of medication management, key components of medication management model and how medication management has been implemented, where relevant. For empirical studies, study design and methodology, study setting, sample size, analytical approach used and main findings will also be extracted. A second reviewer will independently extract data from a sample of studies to ensure consistency in the process.

### Risk of bias

Two independent reviewers will use the Mixed Methods Appraisal Tool (MMAT) version 2018 [22] to critically appraise eligible studies. This tool allows the appraisal of quantitative, qualitative and mixed methods studies. Any disagreements will be resolved by the inclusion of a third independent reviewer and discussion to achieve consensus.

### Data synthesis

For objective 1 (to describe how medication management in H@H is conceptulised) and objective 2 (to describe and identify key components of medication management in H@H), all sections of eligible papers will be read and coded inductively using thematic synthesis [23]. In addition, to meet objective 2, the Systems Engineering Initiative for Patient Safety (SEIPS) 2.0 model [24] will be used to code textual data deductively. The SEIPS 2.0 model is a generic system model that shows the elements of a work system and types of work processes that may be required to produce a range of outcomes for different stakeholders. Analyses based on the SEIPS 2.0 model will add to the inductive analysis, to close any potential gaps in our interpretation of findings reported in eligible papers.

Objective 3 focuses on the complex area of implementation of interventions and in this study, the implementation of medication management services within H@H models globally. Therefore, the Consolidated Framework for Implementation Research (CFIR) will be used to code data deductively from eligible studies [25]. The CFIR constructs includes intervention characteristics (SEIPS 2.0 and the thematic synthesis will describe the components of the intervention i.e. medication management, and therefore different from this construct), outer and inner settings, characteristics of individuals using the intervention and process of implementation.

### Patient and public involvement

The systematic review will be based on published data and no patients or members of the public were/will be involved in the design of the study, interpretation or dissemination of the findings.

## Data Availability

No datasets were generated or analysed during the current study. All relevant data from this study will be made available upon study completion.

## Ethics and dissemination

This systematic review will use secondary data sources from published documents, and as such research ethical approval was not required. We will disseminate the findings of this study in a peer-reviewed journal and national/international conference(s).

## Authors’ contributions

SM, RL and CC conceived the idea for the study. All authors collaborated in designing the study. The protocol was drafted by SM, CC, ZA and RL. All authors contributed to the critical revision of the manuscript. All authors read and approved the final manuscript. RL is the guarantor of the review.

## References

1. McKinsey and Company (2020) The next normal: the future of hospital care: A better patient experience. Available online at https://www.mckinsey.com/~/media/McKinsey/Featured%20Insights/The%20Next%20Normal/The-Next-Normal-Healthcare-collection-2020 Accessed 31/3/22

2. Nuffield Trust (2014) NHS hospitals under pressure: trends in acute activity up to 2022. Available online at https://www.nuffieldtrust.org.uk/files/2017-01/hospitals-under-pressure-web-final.pdf

3. Health Foundation (2021) Our ageing population: how ageing affects health and care need in England. Available online at https://www.health.org.uk/publications/our-ageing-population

4. British Medical Association (2021) NHS hospital beds data analysis. Available online at https://www.bma.org.uk/advice-and-support/nhs-delivery-and-workforce/pressures/nhs-hospital-beds-data-analysis

5. Hamad M and Connolly V (2018) Ambulatory emergency care – improvement by design. Clinical Medicine, 18(1):69–74

6. National Health Service (2019) NHS Long Term Plan. Available at https://www.longtermplan.nhs.uk/online-version/chapter-1-a-new-service-model-for-the-21st-century/2-the-nhs-will-reduce-pressure-on-emergency-hospital-services/ Accessed 31/3/22

7. Melis R, Rikkert M, Paker S and van Eijken M (2004) What is intermediate care? British Medical Journal. 329 (7462) 360–361.

8. The Nuffield Trust (2021) Centralisation of specialist health care services: a mixed-methods programme. Available online https://www.nuffieldtrust.org.uk/project/centralisation-of-specialist-health-care-services-a-mixed-methods-programme. Accessed 22/4/22

9. Statista (2021) Hospital bed density in select countries as of 2019. Available online https://www.statista.com/statistics/283273/oecd-countries--hospital-bed-density/ Accessed 31/3/22

10. Ham 2021. The challenges facing the NHS in England in 2021. British Medical Journal Available online https://www.bmj.com/content/371/bmj.m4973 Accessed 25/2/22

11. Ferrel M and Ryan J (2020) The impact of COVID-19 on medical education. Cureus. 13 (3) e7492

12. Deloitte (2022) 2022 Global Health Care Outlook. Are we finally seeing the long-promised transformation. Available online at https://www2.deloitte.com/content/dam/Deloitte/global/Documents/Life-Sciences-Health-Care/gx-health-care-outlook-Final.pdf Accessed 31/3/22

13. Kings Fund (2021) Health and care in 2021: what can we expect. Available online at https://www.kingsfund.org.uk/blog/2021/01/health-care-2021-what-can-we-expect

14. The Commonwealth Fund (2020) As the pandemic evolves so do ambulatory care practices. Available online https://www.commonwealthfund.org/publications/2020/jun/pandemic-evolves-so-do-ambulatory-care-practices Accessed 31/3/22

15. KPMG (2022) Covid-19: Recovery and resilience in healthcare. Available online https://home.kpmg/xx/en/home/industries/healthcare/covid-19-and-healthcare/covid-19-recovery-and-resilience-healthcare.html Accessed 31/3/22

16. Balatbat C, Kadakia K, Dzau V and Offodile A (2021) No place like home: Hospital at home as a post-pandemic frontier for care delivery innovation. Available online https://catalyst.nejm.org/doi/full/10.1056/CAT.21.0237 Accessed 1/4/22

17. Shepperd S, Butler C, Cradduck-Bamford A et al. (2021) Is comprehensive geriatric assessment admission avoidance hospital at home an alternative to hospital admission for older persons? Annals of Internal Medicine, 174(7): 889–898.

18. UK Hospital at Home society (2022) What is hospital at home? Available online at: https://www.hospitalathome.org.uk/ Accessed 27/11/21

19. NHS England (2021) Guidance note: frailty virtual ward (Hospital at Home for those living with frailty). Available online at: B1207-ii-guidance-note-frailty-virtual-ward.pdf(england.nhs.uk)

20. Centre for Medicare and Medicaid Services (2020) CMS announces comprehensive strategy to enhance Hospital capacity amid Covid-19 surge. Available at https://www.cms.gov/newsroom/press-releases/cms-announces-comprehensive-strategy-enhance-hospital-capacity-amid-covid-19-surge Accessed 27/11/21

21. Moher, D., Shamseer, L., Clarke, M. et al. (2015) Preferred reporting items for systematic review and meta-analysis protocols (PRISMA-P) 2015 statement. Syst Rev 4, 1 (2015). https://doi.org/10.1186/2046-4053-4-1

22. Hong, P Pluye, S Fàbregues, G Bartlett, F Boardman, M Cargo, P Dagenais, M-P Gagnon, F Griffiths, B Nicolau, A O’Cathain, M-C Rousseau, I Vedel (2018). Mixed Methods Appraisal Tool (MMAT), Version 2018 Canadian Intellectual Property Office, Industry, Canada.

23. Thomas J, Harden A. (2008) Methods for the thematic synthesis of qualitative research in systematic reviews. BMC Med Res Methodol.;8(45). https://doi.org/10.1186/1471-2288-8-45.

24. Holden RJ, Carayon P, Gurses AP, et al. (2013) SEIPS 2.0: a human factors framework for studying and improving the work of healthcare professionals and patients. Ergonomics 56:1669–86.

25. Damschroder LJ, Aron D, Keith R, et al. (2009) Fostering implementation of health services research findings into practice: a consolidated framework for advancing implementation science. Implement Sci. 4:50.

